# Clinical phenotypes of uveal melanoma in patients with germline pathogenic/likely pathogenic *BAP1* variants

**DOI:** 10.64898/2026.06.29.26356877

**Authors:** Reham Abdalla, Olivia B. Taylor, Joseph McElroy, Kaylee Ramsey, Lindsey Byrne, Abdelrahman M. Elsayed, Colleen M. Cebulla, Mohamed H. Abdel-Rahman

**Affiliations:** Department of Ophthalmology and Visual Sciences, Havener Eye Institute, The Ohio State University Wexner Medical Center, Columbus, (OH), USA; Department of Ophthalmology, Faculty of Medicine, Assiut University, Assiut, Egypt; Center for Biostatistics, Department of Biomedical Informatics, College of Medicine, The Ohio State University; Division of Human Genetics, Department of Internal Medicine, The Ohio State University Columbus, Columbus, OH, USA; The Ohio State University Comprehensive Cancer Center - Arthur G. James Cancer Hospital and Richard J. Solove Research Institute, Columbus, OH, USA

**Keywords:** *BRCA1-associated protein 1*, *BAP1*, *BAP1* tumor predisposition syndrome, *BAP1*-TPDS, uveal melanoma, Germline BAP1 variants

## Abstract

Germline pathogenic or likely pathogenic variants (GPVs) in *BRCA-1* Associated Protein 1 (*BAP1*) are associated with a spectrum of tumors, including uveal melanoma (UM). Currently, UM patients with *BAP1* GPVs are treated as high-risk class 2 tumors based on mostly empiric data. In the current study, we examined the clinical phenotype of a cohort of 29 UM patients with *BAP1* GPVs. We also carried out a systematic review of the literature of UM patients with *BAP1* GPVs. We observed that UM patients with *BAP1* GPVs have significantly lower median age of diagnosis compared to median age reported in UM patients in the Surveillance, Epidemiology, and End Results Program (SEERS) database. Metastatic risk and overall survival in the UM *BAP1* GPVs cohort were statistically significant from those in patients with class 1 tumors, but were comparable to those observed in UM patients with class 2 tumors. In UM *BAP1* GPVs treated with radiation (n=12), no secondary cancers were observed in the field of radiation in a median 26.5 months (range, 4–119 months) follow up period. One patient experienced a separate growth of UM at a distinct location within the same eye. These data support managing UM in patients with *BAP1* GPVs as aggressive class 2 tumors, following the currently established standard of care for these high-risk tumors.

**Key Points:**

- A small percent of uveal melanomas is attributed to inherited (germline) pathogenic variants in the *BAP1* gene. The clinical outcome of these patients is unclear.
- We studied clinical characteristics, secondary tumors and survival outcomes of uveal melanoma patients with germline *BAP1* pathogenic variants.
- Uveal melanoma patients with germline *BAP1* pathogenic variants were more likely to be diagnosed earlier and their survival outcomes were poorer than those with class 1 tumors, but similar to those with class 2 tumors. In patients treated with radiation, no secondary malignancies were observed but one UM patient had a separate growth of UM at distinct location within the same eye.

## Introduction

Uveal melanoma (UM) is the most common primary intraocular malignancy in adults and remains clinically important because of its strong metastatic potential. Molecular profiling has transformed prognostication by dividing tumors into two major groups, class 1 and class 2, which are determined by gene expression profiling (GEP) or monosomy of chromosome 3.^1,2^ Class 1 and/or disomy 3 tumors generally show a more favorable outcome and class 2 and/or monosomy 3 tumors show a much higher risk of metastasis. This classification has improved risk stratification beyond traditional clinicopathologic features and is now widely used to guide surveillance and counseling.

The only known high-penetrance susceptibility gene for familial UM is *BRCA1-associated Protein 1* (*BAP1*). Approximately 1.6% and 3% of UM patients have been found to harbor germline pathogenic variants (GPVs) in *BAP1*.^3^ GPVs in *BAP1* predispose carriers to a spectrum of tumors, referred to as *BAP1*-tumor predisposition syndrome (*BAP1*-TPDS). The four common cancers observed in BAP1-TPDS include UM, cutaneous melanoma, mesothelioma, and renal cell carcinoma; in addition to the preneoplastic cutaneous *BAP1*-inactivated melanocytic tumors (BIMTs).^4^ UM remains one of the most frequently reported cancers in the *BAP1*-TPDS literature with ~16% of non-probands presenting with UM, although this estimate may be high due to ascertainment bias.^5^

The National Comprehensive Cancer Network (NCCN) acknowledges that GPVs in *BAP1* are a risk factor for developing UM, with larger lesions and aggressive phenotypes. However, it remains to be identified whether germline *BAP1* status should be directly incorporated into risk stratification with molecular class for providers to determine the best management and follow-up frequency.^6^ Gupta *et al*. previously studied the clinical characteristics of a small cohort of eight cases of UM patients with GPVs and reported that tumor diameter, evidence of ciliary body involvement, and incidence of metastasis were significantly higher among patients with *BAP1* GPVs.^7^ However, they observed that tumor diameter, *not* germline *BAP1* status, was the only independent risk factor for metastasis. Thus, a larger cohort of UM patients is needed to determine whether the presence of *BAP1* GPVs significantly impact metastasis and overall survival. This cross-sectional chart review assesses the age of diagnosis and clinical phenotypes in UM patients with *BAP1*-TPDS, including cases from our registry and the published literature, compared with UM patients without germline mutations and population-level UM incidence data from SEER.

## Methods

### Retrospective Chart Review

Retrospective chart review was conducted on uveal melanoma patients enrolled to the Ohio State University (OSU) *BAP1*-TPDS registry “Frequency and Clinical Phenotype of *BAP1* Hereditary Predisposition Syndrome” (NCT04792463 | Registered 3 March 2015 | https://www.clinicaltrials.gov/) or to the OSU Genetics of Uveal Melanoma study. Family members from the *BAP1*-TPDS registry that were determined to be obligate *BAP1* P/LP carriers were included in this cohort. Informed consent was obtained from each patient. The following clinical data were recorded: patient age at diagnosis, sex, personal and family history of cancer, ciliary body involvement, date of earliest detected metastasis, date and cause of death, and date of the last follow-up. Molecular prognostic class (class 1 or class 2) or monosomy of chromosome 3 was determined through testing in a Clinical Laboratory Improvement Amendments (CLIA)–certified clinical laboratories. Germline *BAP1* testing was carried out in CLIA-certified laboratories or at our research laboratory according to published protocols.^8^ Positive results from research testing were confirmed in a CLIA certified laboratory.

Systematic review of English-language peer-reviewed articles was conducted to identify cases of *BAP1* P/LP variants in uveal melanoma patients (Figure 1). Published articles in PubMed and Scopus were reviewed. Variant cDNA change, demographics, tumor characteristics, treatment outcomes, and personal/family history of other cancers were recorded.

**Figure 1:**
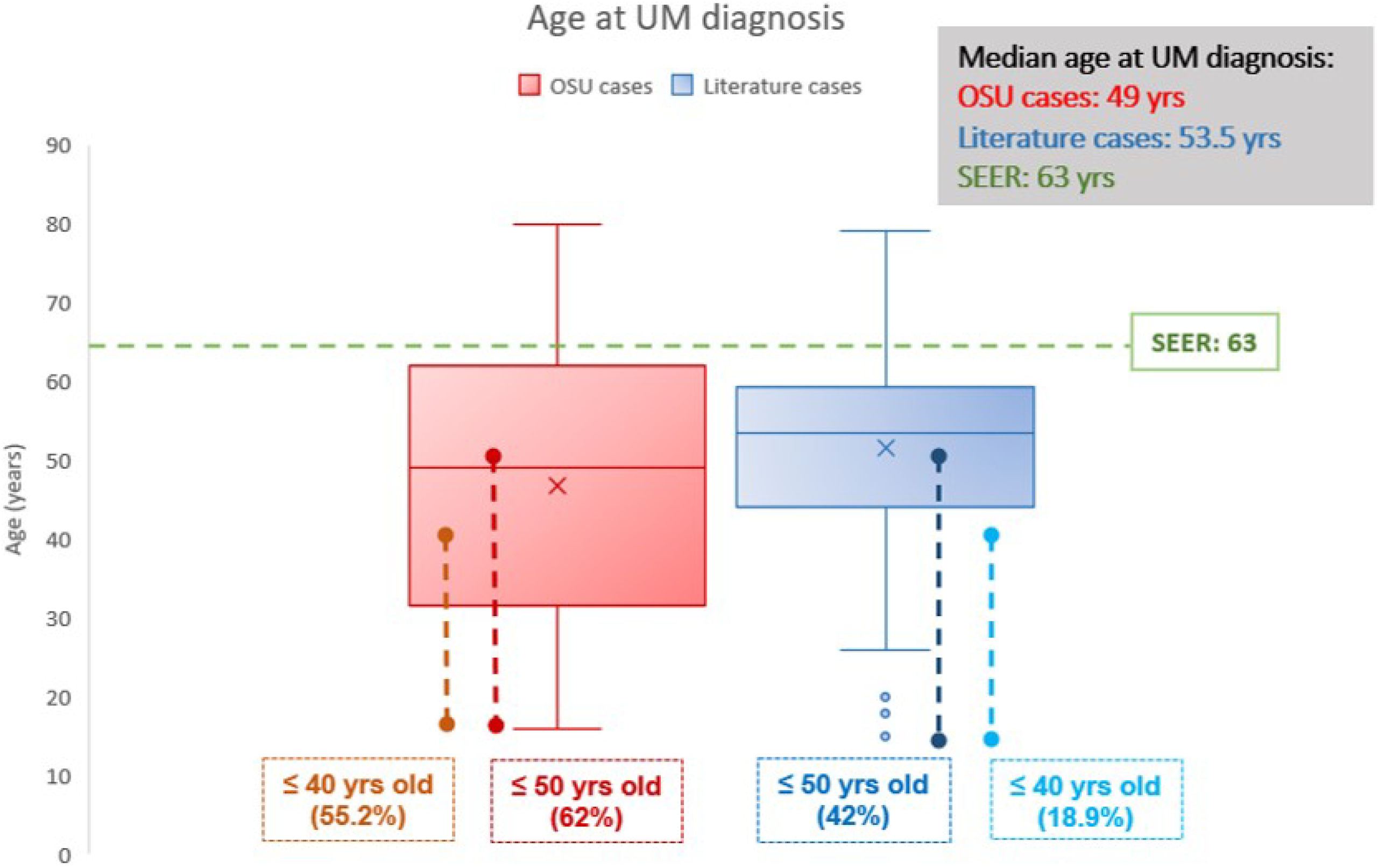
Distribution of age at UM diagnosis among OSU and literature germline BAP1 P/LP cases relative to the SEER-reported median age of diagnosis.

### Statistical Analysis

Significant differences between various demographic and clinical features between BAP1 germline cases and non-germline cases was assessed using student’s t-test and Fisher exact test for continuous and nominal variables, respectively. Kaplan-Meier survival curves with log-rank tests were employed for time to metastasis and overall survival. Cox proportional hazards models were constructed with tumor diameter included as a covariate. For comparisons involving BAP1 germline, class 1, and class 2 tumors, the overall significance of the three level group variable in the Cox model was assessed using ANOVA omnibus tests. A p-value of 0.05 was considered as the significance threshold for this study.

## Results

Patients diagnosed with UM with *BAP1* GPVs were ascertained from IRB-approved studies at Ohio State University (n=29) and from a systematic literature review (n=83) (Figure S1). The median age of diagnosis for OSU cases was 49 years (range: 16-77), ~4.5 years lower than the published literature estimate (53.5 years, range: 17-79) (Table 1). Notably, the SEER-reported median age of UM diagnosis, 63 years, was ~10 and ~14 years higher than the OSU and literature *BAP1*+ cohort, respectively (Table 1, Figure 1).

**Table 1.**
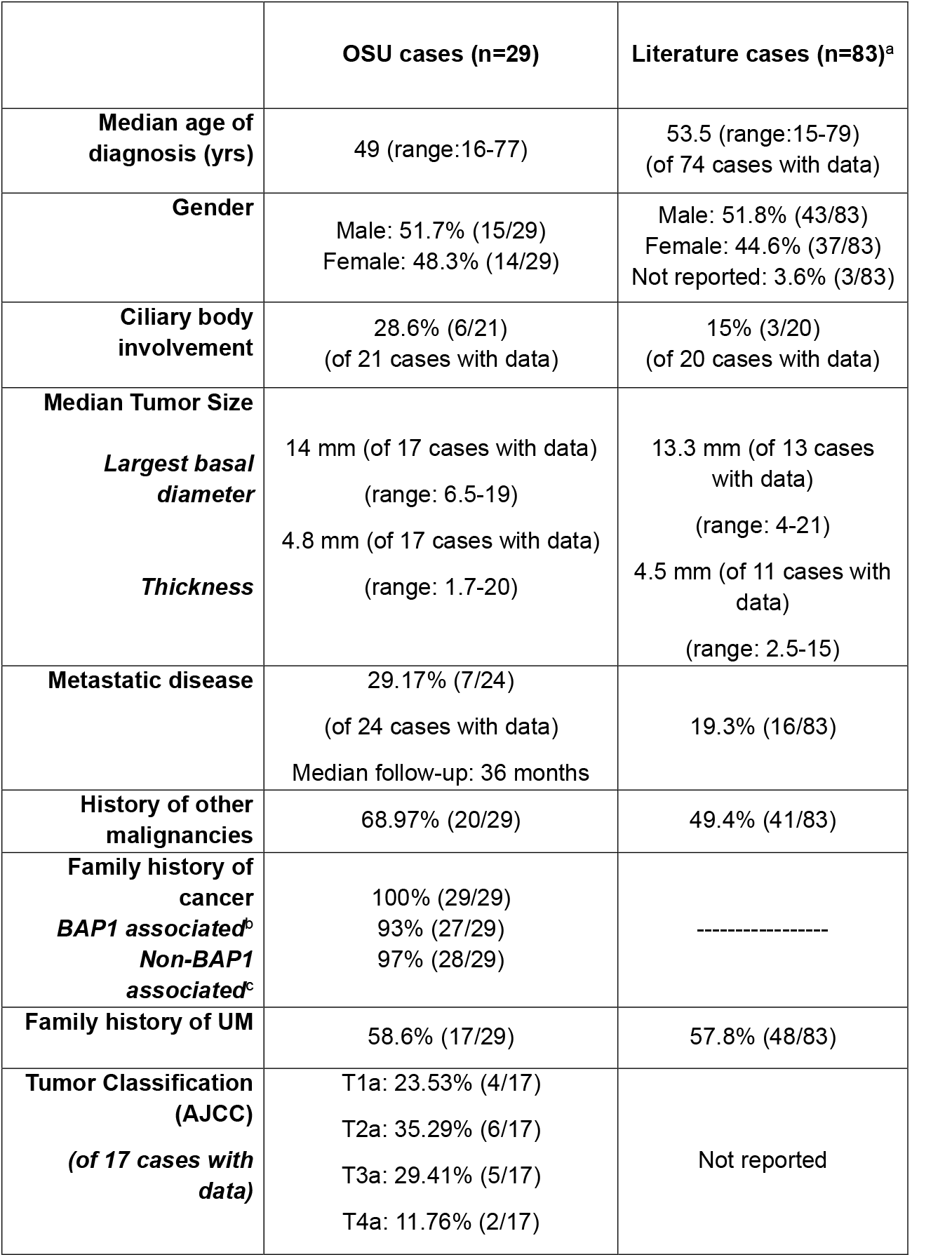
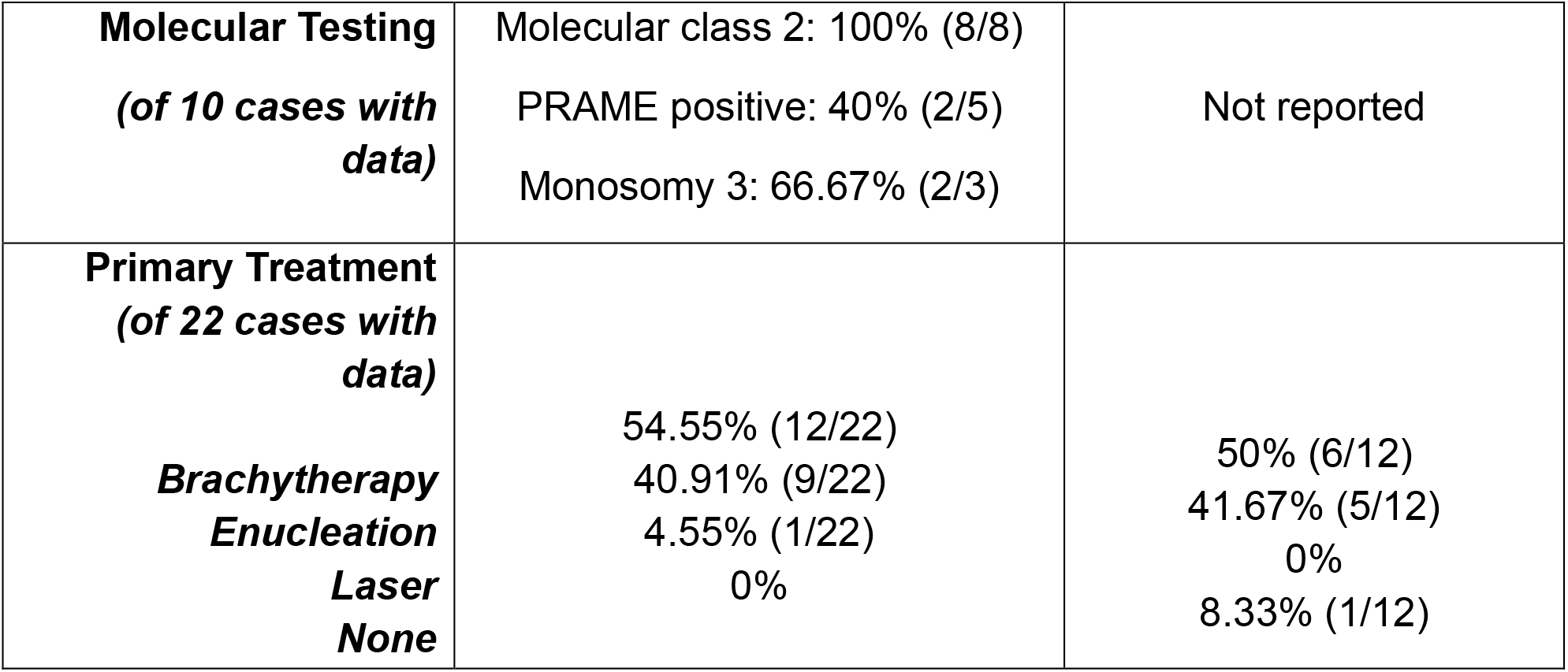
Summary of clinical phenotype of UM patients with germline BAP1 pathogenic variants. Abbreviations: OSU, Ohio State University; BAP1, BRCA1 Associated Protein 1; UM, Uveal Melanoma; AJCC, American Joint Committee on Cancer; PRAME, Preferentially Expressed Antigen in Melanoma ^a^OSU cases were excluded. ^b^BAP1 associated tumors: Basal cell carcinoma, Renal cell carcinoma, Mesothelioma, Cutaneous melanoma and meningioma. ^c^Non-BAP1 associated tumors: Liver, Lung, Breast, Pancreas, Bladder, Colon, Adrenal, Prostate, Testicular, Ovarian, SCC, Leukemia, Lymphoma and Brain

|  | OSU cases (n=29) | Literature cases (n=83)<sup>a</sup> |
| --- | --- | --- |
| **Median age of diagnosis (yrs)** | 49 (range:16-77) | 53.5 (range:15-79)<br>(of 74 cases with data) |
| **Gender** | Male: 51.7% (15/29)<br>Female: 48.3% (14/29) | Male: 51.8% (43/83)<br>Female: 44.6% (37/83)<br>Not reported: 3.6% (3/83) |
| **Ciliary body involvement** | 28.6% (6/21)<br>(of 21 cases with data) | 15% (3/20)<br>(of 20 cases with data) |
| **Median Tumor Size** |  |  |
| ***Largest basal diameter*** | 14 mm (of 17 cases with data)<br>(range: 6.5-19) | 13.3 mm (of 13 cases with data)<br>(range: 4-21) |
| ***Thickness*** | 4.8 mm (of 17 cases with data)<br>(range: 1.7-20) | 4.5 mm (of 11 cases with data)<br>(range: 2.5-15) |
| **Metastatic disease** | 29.17% (7/24)<br>(of 24 cases with data)<br>Median follow-up: 36 months | 19.3% (16/83) |
| **History of other malignancies** | 68.97% (20/29) | 49.4% (41/83) |
| **Family history of cancer**<br>***BAP1 associated<sup>b</sup>***<br>***Non-BAP1 associated<sup>c</sup>*** | 100% (29/29)<br>93% (27/29)<br>97% (28/29) | ----- |
| **Family history of UM** | 58.6% (17/29) | 57.8% (48/83) |
| **Tumor Classification (AJCC)**<br>***(of 17 cases with data)*** | T1a: 23.53% (4/17)<br>T2a: 35.29% (6/17)<br>T3a: 29.41% (5/17)<br>T4a: 11.76% (2/17) | Not reported |
| **Molecular Testing**<br>*(of 10 cases with data)* | Molecular class 2: 100% (8/8)<br>PRAME positive: 40% (2/5)<br>Monosomy 3: 66.67% (2/3) | Not reported |
| **Primary Treatment**<br>*(of 22 cases with data)*<br>*Brachytherapy*<br>*Enucleation*<br>*Laser*<br>*None* | 54.55% (12/22)<br>40.91% (9/22)<br>4.55% (1/22)<br>0% | 50% (6/12)<br>41.67% (5/12)<br>0%<br>8.33% (1/12) |

In the OSU cohort, 93% (27/29) of patients reported having a family history of *BAP1*-associated cancers^9^, and 68.97% (20/29) of patients had a personal history of other malignancies in addition to uveal melanoma (Table 1). In the published literature, a personal history of other malignancies was observed in 95% (39/41 cases with reported family history). Many patients with personal cancer/tumor history had another *BAP1*-associated malignancy, including basal cell carcinoma, renal cell carcinoma, mesothelioma, skin melanoma, and meningioma (Table S1).

Median tumor dimensions (diameter and thickness) were similar between OSU and published cases with available clinical data (Table 1). In the OSU cohort, seventeen tumors were given an AJCC tumor classification; a majority were classified as T2a (35.3%, 6/17) or T3a (29.4%, 5/17) tumors. Because many patients were accrued 10-14 years ago, only a small number (n=10) of OSU patients had any type of clinical laboratory molecular tumor testing. Of these, all were identified as class 2 tumors, 40% (2/5) of tumors were PRAME positive, and 66.67% (2/3) showed chromosome 3 monosomy. Molecular testing information was not available from the published literature. Among patients treated with radiation (n=12) no patients developed secondary malignancies but one patient developed two distinct local secondary UM growths observed at 84 months follow-up in the same eye.

Tumor characteristics of OSU *BAP1*-TPDS cases were compared to UM patients who had exome sequencing and were found to have no GPVs in BAP1 (Table 2). Non-germline cases were stratified by class 1 (better prognosis) and class 2 (aggressive) tumors. Consistent with the comparisons to the SEER estimated median age, *BAP1*-TPDS patients had significantly lower age of diagnosis relative to class 1 (p<0.001) and class 2 non-germline patients (p<0.001). There were no significant sex differences or differences in ciliary body involvement of the tumor. Tumor size was significantly greater in class 2 tumors relative to class 1 (p<0.005). Notably, germline *BAP1* patients had a metastatic risk comparable (i.e. non-significant) to that of class 2 UM and significantly higher than that of class 1 UM (p<0.05).

**Table 2.** Clinical characteristics of uveal melanomas in the studied cohort stratified by molecular class 1 and 2 tumors and germline BAP1 mutation status. ^a^P-values determined by Fisher exact test Abbreviations: BAP1, BRCA1 Associated Protein 1; OSU, Ohio State University

|  | OSU BAP1 germline cases<br>(n=29) | Molecular Class 1 (n=93) | Molecular Class 2<br>(n=31) | <sup>a</sup>p- values |  |  |
| --- | --- | --- | --- | --- | --- | --- |
|  |  |  |  | *Class 1*<br>x<br>*Class 2* | *BAP1 germline*<br>x<br>*Class 1* | *BAP1 germline*<br>x<br>*Class 2* |
| **Median age of diagnosis**<br>**(years)** | 49 (range:16-77) | 60 (range:18-85) | 66 (range:31-84) | 0.122666368 | **0.000111013** | **0.000189613** |
| **Gender**<br>*Male*<br>*Female* | 51.7% (15/29)<br>48.3% (14/29) | 44.1% (41/93)<br>55.9% (52/93) | 54.8% (17/31)<br>45.2% (14/31) | 0.406203 | 0.525704 | 1 |
| **Ciliary body involvement** | 28.6% (6/21)<br>(of 21 cases with data) | 17.2% (16/93) | 13.3% (4/30)<br>(of 30 cases with data) | 0.779193 | 0.234563 | 0.283032 |
| **Median Tumor Size**<br>*Median largest basal*<br>*diameter*<br>*Diameter > 12 mm* | 14 mm<br>(of 17 cases with data)<br>(range: 6.5-19)<br>58.8% (10/17) | 11.9 mm<br>(of 91 cases with data)<br>(range: 3.1-19)<br>44.0% (40/91) | 15 mm<br>(of 27 cases with data)<br>(range: 10-19.4)<br>66.7% (18/27) | **0.002026339**<br>1 | 0.127865807<br>0.297541 | 0.382243375<br>0.749451 |
| **Metastatic disease** | 29.17% (7/24)<br>(of 24 cases with data) | 8.7% (8/92)<br>(of 92 cases with data) | 41.9% (13/31) | **0.000088** | **0.014420** | 0.403043 |

UM patients with *BAP1* GPVs demonstrated metastasis-free survival and overall survival outcomes which were statistically similar to patients with class 2 tumors, with no significant difference before or after adjustment for tumor diameter (Table 3, Figure 2). Conversely, germline *BAP1* patients showed significantly shorter metastasis-free survival (p<0.005) and overall survival (p<0.05) than molecular class 1 tumors; this effect persisted after adjustment for continuous tumor diameter. However, this association was attenuated, losing statistical significance, when tumor diameter was included as a covariate (dichotomized to the clinically relevant threshold, >12 mm), suggesting that the increased prevalence of large UM tumors in *BAP1*-TPDS patients could partially contribute to the adverse clinical outcomes observed in this cohort.

**Table 3:** Cox proportional hazards analysis of time to metastasis and overall survival between germline BAP1 P/LP patients and non-germline patients with molecular class 1 or 2 tumors. Abbreviations: BAP1, BRCA1-Associated Protein 1; CI, Confidence Interval

|  | Comparison | Hazard Ratio (95% CI) | p-value | Continuous diameter significant? | Diameter threshold (> 12 mm) significant? |
| --- | --- | --- | --- | --- | --- |
|  | Class 1 vs Class 2 | 8.357 (2.551-27.374) | **0.0004531** | **yes** | no |
| ***Time to metastasis*** | Germline BAP1 vs Class 1 | 7.416 (1.972-27.909) | **0.003038** | **yes** | no |
|  | Germline BAP1 vs Class 2 | 1.06 (0.349-3.22) | 0.918 | no | no |
| ***Overall survival*** | Class 1 vs Class 2 | 2.241 (1.287-6.467) | 0.1354 | no | no |
|  | Germline BAP1 vs Class 1 | 3.621 (1.086-12.076) | **0.03622** | **yes** | no |
|  | Germline BAP1 vs Class 2 | 1.549 (0.435-5.513) | 0.4992 | no | no |

**Figure 2:**
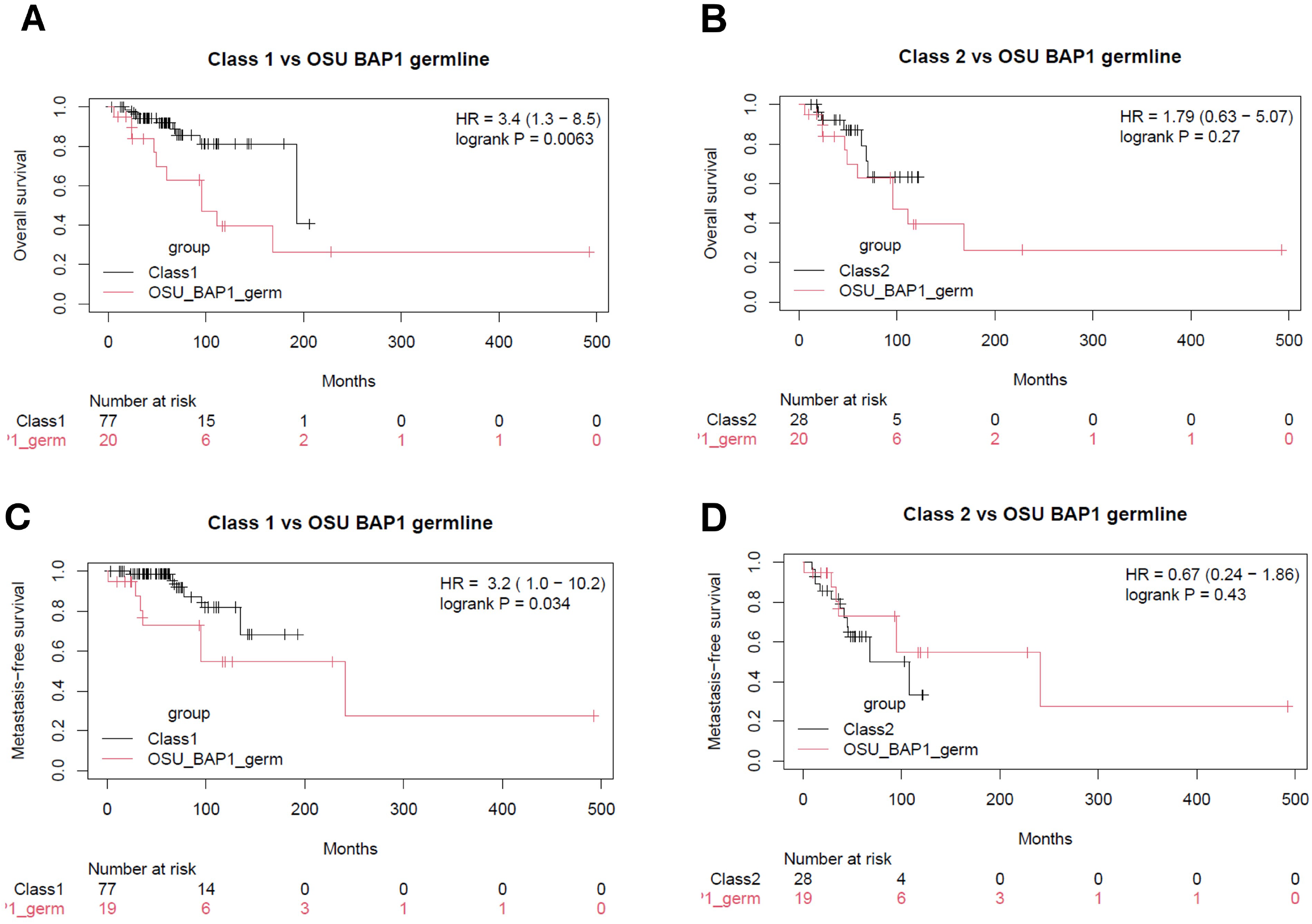
Germline mutations in BAP1 are associated with poor survival outcomes in uveal melanoma patients. Kaplan–Meier curves comparing overall survival (A, B) and metastasis-free survival (C, D) between uveal melanoma patients with germline BAP1 mutations and either Class 1 or Class 2. Germline BAP1 cases showed significantly poor overall survival and metastasis-free survival compared with Class 1 tumors (A: HR = 3.4, log-rank p = 0.0063; C: HR = 3.2, log-rank p = 0.034), but outcomes were not significantly different from Class 2 tumors (B: HR = 1.79, p = 0.27; D: HR = 0.67, p = 0.43). Tick marks indicate censored observations, and numbers at risk are shown below each plot.

## Discussion

This study expands the phenotypic characterization of UM patients with *BAP1* GPVs by combining a contemporary institutional cohort with previously published cases. Overall, our findings reinforce the concept that UM from patients with BAP1 GPVs represents a distinct clinical entity characterized by earlier age at diagnosis, frequent personal and family histories of cancer, high prevalence of tumors associated with *BAP1* tumor predisposition syndrome (*BAP1*-TPDS), and more aggressive UM tumor characteristics. These observations are consistent with the growing body of literature demonstrating that inherited *BAP1* pathogenic variants confer susceptibility to a broad spectrum of malignancies with varying tumor pathology and should prompt consideration of genetic evaluation and long-term surveillance in affected individuals and their relatives.

The demographic characteristics of our cohort were similar to those reported previously.^5,7,10^ Male and female patients were nearly equally represented in both the OSU and literature cohorts, suggesting that germline *BAP1* variants do not substantially alter the sex distribution observed in UM overall. One of the most striking findings of this study is the substantially younger age at diagnosis among patients with *BAP1* GPVs compared with the general UM population. The median age at diagnosis in the OSU cohort was 49 years compared with 53.5 years among published cases and 63 years in the SEER population.^11^ Over half of the OSU patients and one fifth of the literature cases were diagnosed by age 40. Moreover, age of onset in the OSU *BAP1* GPVs cohort was significantly lower than patients with no *BAP1* germline mutations who had class 1 and 2 tumors. These findings support previous reports demonstrating that UM develops approximately one decade earlier in germline *BAP1* carriers than in sporadic cases supporting early ophthalmic surveillance.^10^ Earlier disease onset likely reflects constitutional loss of one *BAP1* allele, requiring only a second somatic event for tumor development according to the classical two-hit tumor suppressor model.^10^

There were no significant differences in metastasis-free and overall survival between UM patients with *BAP1s* GPV and molecular class 2 patients with no germline mutations, supporting the management of UM in *BAP1*-TPDS as class 2 aggressive tumors. However, germline *BAP1* tumors are more likely to exceed 12 mm in tumor diameter, which may partially mediate or confound the association between germline *BAP1* status and poorer prognosis. This was a similar outcome to the study conducted by Gupta *et al* in which the effect of *BAP1* mutation status on metastasis was dependent on tumor diameter.^7^ This group assessed continuous diameter which approached significance. In our study, we examined diameter as a continuous measurement or as threshold (>12 mm). When tumor diameter was modeled as a continuous covariate, germline *BAP1* status remained independently associated with worse prognosis, but this association was attenuated when tumor diameter was dichotomized. Due to the size of this cohort, dichotomizing this variable reduces statistical power, removing potentially important within-group variation. Continued long-term follow-up in robustly powered cohorts is needed to more rigorously tease out the independent effects of germline *BAP1* mutations on clinical outcomes.

Given the association of *BAP1* mutations with DNA damage repair defects, the utility and potential complications of radiation in patients with *BAP1* GPVs are largely unknown. Patients with GPVs in other tumor suppressor genes, including *TP53, RB1*, and *NF1*, have been found to be at higher risk of radiation-induced secondary malignancies occurring several years after treatment.^12–15^ Notably, secondary malignancies were found to be a major complication of external beam radiation therapy for treatment of retinoblastoma in patients with *RB1* GPVs, leading to its discontinuation as a first-line therapy.^16–18^ These secondary malignancies, mainly soft tissue and bone sarcoma, may be diagnosed more than 10 years after radiation therapy.^19^ To the authors’ knowledge, only two literature cases of radiation-induced malignancies following radiation for UM have been reported. The first reported an osteosarcoma appearing at the site of radiation nine years after orbital apex radiotherapy.^20^ The second proposed that a meningioma appearing in the cavernous sinus 9 following adjuvant proton beam therapy was an effect of the patient’s UM treatment.^21^ No secondary cancers are reported in the literature in UM patients with *BAP1* GPVs treated with radiation. No secondary non-UM cancers were found in the field of radiation in the OSU patient cohort (data not shown). However, given the relatively short follow up of these patients (median: 26.5 months, range: 4-119 months), the long-term effect of radiation is unknown. It remains to be determined whether localized radiation through plaque brachytherapy or external beam radiation can cause secondary malignancies among patients with *BAP1* GPVs.

This study has several limitations. In addition to the retrospective design, missing clinical and molecular data in published reports limited direct comparisons across cohorts, while differences in follow-up duration likely influenced estimates of metastatic disease. In addition, the relatively small institutional cohort reduces statistical power for subgroup analyses. Despite these limitations, combining institutional experience with published cases provides one of the more comprehensive clinical descriptions of UM with *BAP1* GPVs and highlights consistent phenotypic patterns across independent populations.

## Conclusions

Germline pathogenic *BAP1* variants define a distinct subset of uveal melanoma, characterized by earlier onset, a broad spectrum of personal and familial cancer history, and poorer prognosis. Importantly, patients with *BAP1*-TPDS have equivalent metastatic risk and overall survival outcomes to molecular class 2 tumors with no underlying germline mutation, supporting the treatment of these patients as a similarly high-risk population. Larger, perhaps multi-center, studies with extended follow-up are warranted to further define the prognostic impact of underlying germline *BAP1* mutations and utility/complications of radiation in these patients.

## Author Contributions

*Concept and design:* RA, CC, MHA

*Acquisition, analysis, or interpretation of data*: RA, OT, KR

*Drafting of the manuscript:* RA, OT

*Critical review of the manuscript for important intellectual content:* RA, OT, MHA, JM, CC, KR, LB, AME

*Statistical analysis:* JM, OT, RA

*Obtained funding:* MHA, LB, JM, CC

*Administrative, technical, or material support:* CC, MHA

*Supervision:* CC, MHA

## Ethics Statement

The ethics committee/IRB of Ohio State University gave ethical approval for this work. Written informed consent was obtained for all prospectively enrolled participants, and a waiver of informed consent was obtained for retrospective chart review components.

## Data Availability

All original data generated in support of this research will be made available at reasonable request to the corresponding author.

---

**Table 1S.** Percentage of other primary malignancies in UM patients with *BAP1* germline pathogenic variants. Abbreviations: BAP1, BRCA1 Associated Protein 1; OSU, Ohio State University ^a^Other cancers include Liver, Pancreas, Prostate, SCC, Thymus and Brain

| **BAP1 associated tumors:** | **OSU cases (n=20)** | **Literature cases (n=41)** |
| --- | --- | --- |
| **Confirmed association** | **Total: 60% (12/20)** | **Total: 95% (39/41)** |
| Basal cell carcinoma | 20% (4/20) | 19.5% (8/41) |
| Renal cell carcinoma | 15% (3/20) | 26.8% (11/41) |
| Mesothelioma | 10% (2/20) | 29.3% (12/41) |
| Cutaneous melanoma | 10% (2/20) | 12.2% (5/41) |
| Meningioma | 5% (1/20) | 7.3% (3/41) |
| **Unconfirmed association** | **Total: 20% (4/20)** | **Total: 22% (9/41)** |
| Bladder | 5% (1/20) | 2.4% (1/41) |
| Colon | 5% (1/20) | 4.9% (2/41) |
| Lung | 5% (1/20) | 9.8% (4/41) |
| Breast | 5% (1/20) | 4.9% (2/41) |
| **Other malignancies<sup>a</sup>** | **20% (4/20)** | **15% (6/41)** |

**Figure S1:**
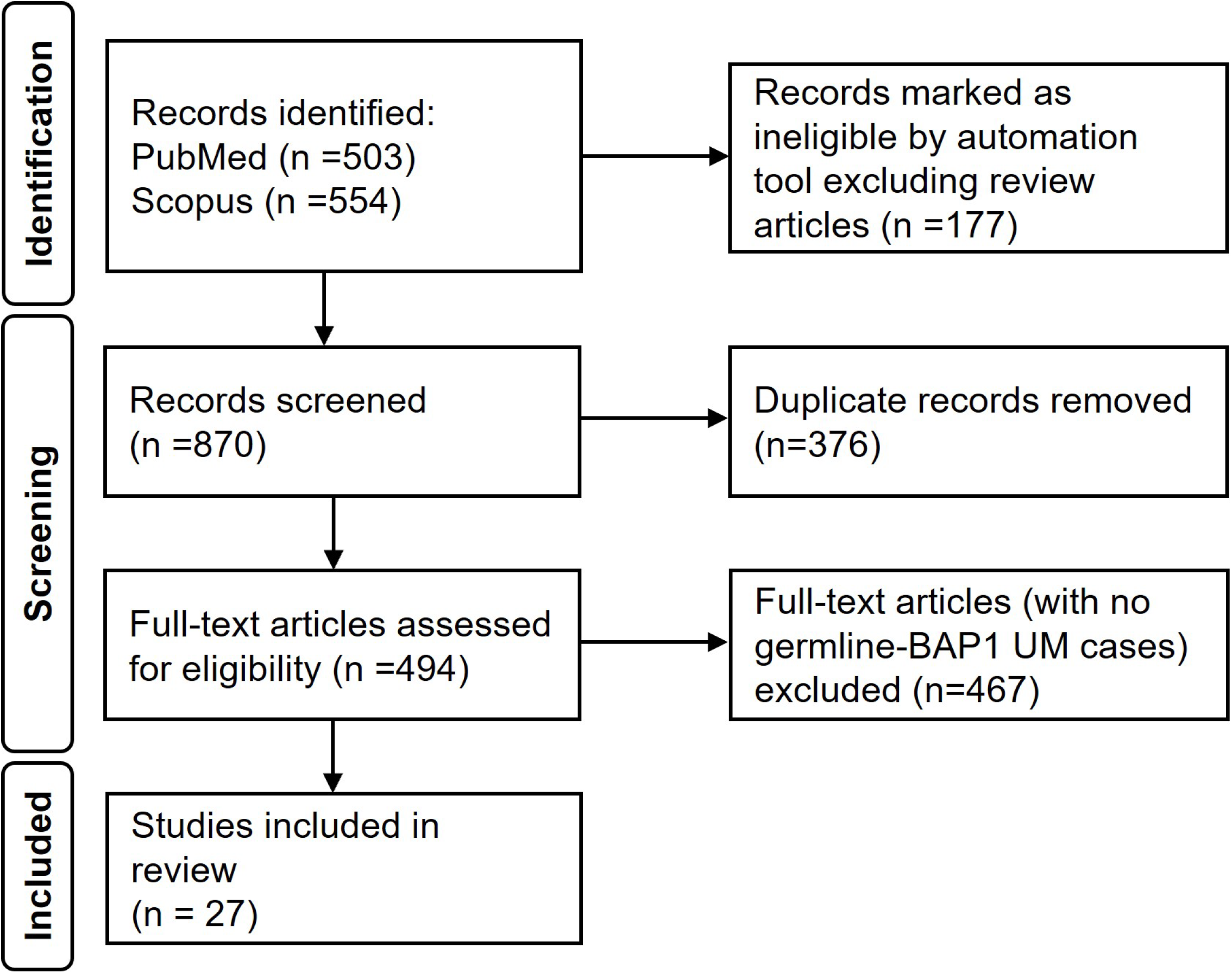
PRISMA flow diagram of systematic literature review for reported germline *BAP1* cases among patients with uveal melanoma.

## References

1. Onken MD, Worley LA, Ehlers JP, Harbour JW. Gene Expression Profiling in Uveal Melanoma Reveals Two Molecular Classes and Predicts Metastatic Death. Cancer Res. 2004;64(20):7205–7209. doi:10.1158/0008-5472.CAN-04-1750

2. Walter SD, Chao DL, Feuer W, Schiffman J, Char DH, Harbour JW. Prognostic Implications of Tumor Diameter in Association With Gene Expression Profile for Uveal Melanoma. JAMA Ophthalmol. 2016;134(7):734–740. doi:10.1001/jamaophthalmol.2016.0913

3. Helgadottir H, Höiom V. The genetics of uveal melanoma: current insights. Appl Clin Genet. 2016;9:147–155. doi:10.2147/TACG.S69210

4. Pilarski R, Byrne L, Carlo MI, Hanson H, Cebulla C, Abdel-Rahman M. BAP1 Tumor Predisposition Syndrome. In: Adam MP, Bick S, Mirzaa GM, Pagon RA, Wallace SE, Amemiya A, eds. GeneReviews®. University of Washington, Seattle; 1993. Accessed June 26, 2026. http://www.ncbi.nlm.nih.gov/books/NBK390611/

5. Walpole S, Pritchard AL, Cebulla CM, et al. Comprehensive Study of the Clinical Phenotype of Germline BAP1 Variant-Carrying Families Worldwide. J Natl Cancer Inst. 2018;110(12):1328–1341. doi:10.1093/jnci/djy171

6. NCCN Guidelines, Melanoma: Uveal. 11 2025. Accessed September 18, 2025. https://www.nccn.org/professionals/physician_gls/pdf/uveal.pdf

7. Gupta MP, Lane AM, DeAngelis MM, et al. Clinical Characteristics of Uveal Melanoma in Patients With Germline BAP1 Mutations. JAMA Ophthalmol. 2015;133(8):881–887. doi:10.1001/jamaophthalmol.2015.1119

8. Boru G, Grosel T, Pilarski R, et al. Germline large deletion of BAP1 and decreased expression in non-tumor choroid in uveal melanoma patients with high risk for inherited cancer. Genes Chromosomes and Cancer. 2019;58(9):650–656. doi:10.1002/gcc.22752

9. Pilarski R, Byrne L, Carlo MI, Hanson H, Cebulla C, Abdel-Rahman M. BAP1 Tumor Predisposition Syndrome. In: Adam MP, Feldman J, Mirzaa GM, Pagon RA, Wallace SE, Amemiya A, eds. GeneReviews®. University of Washington, Seattle; 1993. Accessed July 28, 2025. http://www.ncbi.nlm.nih.gov/books/NBK390611/

10. Singh N, Singh R, Bowen RC, Abdel-Rahman MH, Singh AD. Uveal Melanoma in BAP1 Tumor Predisposition Syndrome: Estimation of Risk. Am J Ophthalmol. 2021;224:172–177. doi:10.1016/j.ajo.2020.12.005

11. Weinberger Y, Bena J, Singh AD. Uveal Melanoma: 5-Year Update on Incidence, Treatment, and Survival (SEER 1975–2020). Ocul Oncol Pathol. 2025;11(1):30–36. doi:10.1159/000543151

12. Miao R, Wang H, Jacobson A, et al. Radiation-induced and neurofibromatosis-associated malignant peripheral nerve sheath tumors (MPNST) have worse outcomes than sporadic MPNST. Radiother Oncol. 2019;137:61–70. doi:10.1016/j.radonc.2019.03.015

13. Dutra MP, Rodrigues CM, Peretz-Soroka H, et al. Radiation-induced sarcomas following childhood cancer - A Canadian Sarcoma Research and Clinical Collaboration Study (CanSaRCC). Cancer Rep (Hoboken). 2023;6(6):e1834. doi:10.1002/cnr2.1834

14. Hendrickson PG, Luo Y, Kohlmann W, et al. Radiation therapy and secondary malignancy in Li-Fraumeni syndrome: A hereditary cancer registry study. Cancer Med. 2020;9(21):7954–7963. doi:10.1002/cam4.3427

15. Casey A, Barliana JD. The incidence of secondary neoplasms in retinoblastoma survivors who underwent radiation therapy: A systematic review and meta-analysis. Taiwan J Ophthalmol. 2025;15(1):45–54. doi:10.4103/tjo.TJO-D-23-00086

16. Mohney BG, Robertson DM, Schomberg PJ, Hodge DO. Second nonocular tumors in survivors of heritable retinoblastoma and prior radiation therapy. Am J Ophthalmol. 1998;126(2):269–277. doi:10.1016/s0002-9394(98)00146-9

17. Fletcher O, Easton D, Anderson K, Gilham C, Jay M, Peto J. Lifetime risks of common cancers among retinoblastoma survivors. J Natl Cancer Inst. 2004;96(5):357–363. doi:10.1093/jnci/djh058

18. Fontanesi J, Parham DM, Pratt C, Meyer D. Second malignant neoplasms in children with retinoblastoma: the St. Jude Children’s Research Hospital experience. Ophthalmic Genet. 1995;16(3):105–108. doi:10.3109/13816819509059968

19. Casey A, Barliana JD. The incidence of secondary neoplasms in retinoblastoma survivors who underwent radiation therapy: A systematic review and meta-analysis. Taiwan Journal of Ophthalmology. 2025;15(1):45. doi:10.4103/tjo.TJO-D-23-00086

20. Morris B, Williams W, Shuttleworth GN. Osteosarcoma after external beam radiation therapy for recurrent choroidal melanoma. Ophthalmic Plast Reconstr Surg. 2006;22(4):301–302. doi:10.1097/01.iop.0000225423.00759.26

21. Scaringi C, Minniti G, Bozzao A, et al. Radiation-induced malignant meningioma following proton beam therapy for a choroidal melanoma. Journal of Clinical Neuroscience. 2015;22(6):1036–1037. doi:10.1016/j.jocn.2014.12.021

